# Early COVID-19 Therapy with Azithromycin Plus Nitazoxanide, Ivermectin or Hydroxychloroquine in Outpatient Settings Significantly Reduced Symptoms Compared to Known Outcomes in Untreated Patients

**DOI:** 10.1101/2020.10.31.20223883

**Authors:** Flavio A. Cadegiani, Andy Goren, Carlos G. Wambier, John McCoy

**Affiliations:** Corpometria Institute, Brasília, DF, Brazil; Applied Biology Inc., Irvine, CA, USA; Brown University, RI, USA

**Keywords:** COVID-19, SARS-CoV-2, hydroxychloroquine, ivermectin, nitazoxanide, antiandrogen, dutasteride, spironolactone, proxalutamide

## Abstract

**Background:** While there was a lack of pharmacological interventions proven to be effective in early, outpatient settings for COVID-19, in a prospective, open-label observational study (pre-AndroCoV Trial) the use of nitazoxanide, ivermectin and hydroxychloroquine demonstrated similar effects, and apparent improvement of outcomes compared to untreated patients. The unexpected apparent positive results led to ethical questions on the employment of further full placebo-control studies in early stage COVID-19. The objective of the present study was to elucidate whether the conduction of a full placebo-control RCT was still ethically viable, through a comparative analysis with two control-groups.

**Materials and methods:** Active group (AG) consisted of mild-to-moderate early stage COVID-19 patients enrolled in the Pre AndroCoV-Trial, treated with nitazoxanide ivermectin, or hydroxychloroquine in selected cases, in association with azithromycin. Vitamin D, vitamin C, zinc, glucocorticoids and anticoagulants, when clinically recommended. Control Group 1 (CG1) consisted of a retrospectively obtained group of untreated patients from the same population as those from the Pre-AndroCoV Trial, and Control Group 2 (CG2) resulted from a precise prediction of clinical outcomes, based on a thorough and structured review of articles indexed in PubMed and MEDLINE and statements by official government agencies and specific medical societies. For both CGs, patients were matched for proportion between sex, age, obesity and other comorbidities. Results: Compared to CG1 and CG2, AG showed a reduction of 31.5 to 36.5% in viral shedding (p < 0.0001), 70 to 85% and 70 to 73% in duration of COVID-19 clinical symptoms when including and not including anosmia and ageusia, respectively ((p < 0.0001 for both), and 100% in respiratory complications through the parameters of the Brescia COVID-19 Respiratory Scale (p < 0.0001). For every 1,000 confirmed cases for COVID-19, a minimum of 140 patients were prevented from hospitalization (p < 0.0001), 50 from mechanical ventilation, and five deaths, when comparing to age-, sex- and comorbidity-matched non-treated patients with similar initial disease severity at the moment of diagnosis.

**Conclusion:** Apparent benefits of the combination between early detection and early pharmacological approaches for COVID-19 demonstrated to be consistent when when compared to different control groups of untreated patients. The potential benefits could allow a large number of patients prevented from hospitalizations, deaths and persistent symptoms after COVID-19 remission. The potential impact on COVID-19 disease course and numbers of negative outcomes and the well-established safety profile of the drugs proposed by the Pre-AndroCoV Trial led to ethical questions regarding the conduction of further placebo control randomized clinical trials (RCTs) for early COVID-19. Early pharmacological approaches including azithromycin in combination with any of the options between nitazoxanide, ivermectin or optionally hydroxychloroquine should be considered for those diagnosed with COVID-19 presenting less than seven days of symptoms. Of the three drugs, we opted for nitazoxanide, due to more extensive demonstration of *in vitro* and *in vivo* antiviral activity, proven efficacy against other viruses in humans, and steadier safety profile.

## Background

Coronavirus Disease 2019 (COVID-19) is a highly heterogeneous and multi-systemic infection caused by the novel Severe Acute Respiratory Syndrome Coronavirus 2 (SARS-CoV-2), first described in November 2019 in Wuhan, China, and further spread worldwide, leading to a non-ceasing pandemic since March 2020 (1-3). The key reasons that may justify the inability to contain its dissemination are based on the some of the specific properties of SARS-CoV-2, including prolonged incubation and viral shedding, large percentage of non-symptomatic (asymptomatic or pre-symptomatic) subjects actively transmitting SARS-CoV-2, persistent survival in surfaces, and highly spreader environments (1,2). In addition, its transmission and dissemination patterns are yet to be fully elucidated (2,3).

Natural course of COVID-19 includes dry couch and rhinorrhea as the prodromic symptoms, possibly indicating direct viral infection by the oral and nasal mucosa retrospectively, followed by feverish, chills, arthralgia, muscle soreness, fatigue, gastrointestinal and upper respiratory tract-related symptoms, and finally followed by fever, anosmia, ageusia and shortness of brief, later during the first stage. Risk factors include aging (above 60 y/o), uncontrolled diabetes, hypertension, obesity, and hyperandrogenic state, in both males and females (4-11).

The current understanding of COVID-19 natural course allows the disease to be divided into three stages. The first stage corresponds to the viral replication and has an approximate duration of seven to ten days. The first stage progressively progresses to a second stage, when inflammatory, immunologic and vascular overreactions occur, eventually leading to a diffuse lung injury, the third stage. The second and third stages are highly variable among individuals and are the actual drivers of COVID-19 severity.

The identification of effective treatments to improve COVID-19 clinical outcomes, mortality and post-COVID manifestations is highly desired while definitive solutions like effective and safe vaccines are not universally available. Targets that address SARS-CoV-2 mechanisms of infection and risk factors allow proposals of more precise therapies to be potentially effective against COVID-19. From the understanding of COVID-19, pharmacological interventions during the first stage are likely the most efficient timing to prevent complications triggered by SARS-CoV-2 and the most relevant window of opportunity to antagonize SARS-CoV-2 infectivity. Direct or indirect antiviral strategies include acute inhibition of ACE2 attached to cell surfaces, the only direct mechanism of SARS-CoV-2 cell entry, increase of circulating ACE2, since it may preclude viral infectivity by coupling with SARS-CoV-2, preventing its entry into cells, and inhibition of transmembrane serine protease 2 (TMPRSS-2), a critical protein that facilitates viral entry through ACE2 (12-21). Repurposing existing drugs may present some advantages over novel molecules, including well-established safety profile, known risks and contraindications, familiarity among health providers, and favorable cost-effectiveness, in case efficacy for COVID-19 is demonstrated (2;22-29).

Controversies regarding treatment during early (first stage) COVID-19 include supposed lack of a pharmacological option with clear benefits. Hydroxychloroquine (HCQ), nitazoxanide (NIT) and ivermectin (IVE), in association with azithromycin (AZI), are popular drugs largely used as *off label* therapies for COVID-19, that have demonstrated *in vitro* antiviral activity and preliminary observational reports as being beneficial against COVID-19, when used until seven days after beginning of symptoms, before respiratory complications and hospitalization (26-40). Use of any of these drugs further in the disease, during second and third stages, have not demonstrated benefits or conflicting findings (2;30).

Antiandrogens could play a protective action against COVID-19 by the inhibition of TMPRSS-2 expression, that finds in androgens its only known modulators (2;9-11;41-43). Indeed, chronic dutasteride users have demonstrated to protect against severe COVID-19 in a variety of male populations (44-46), which encourages the employment of antiandrogens in clinical trials for early COVID-19.

The present group hypothesized that none of the most popular drugs, including HCQ, NIT and IVE, would result in actual benefits in any stage of COVID-19. While we were designing our randomized clinical trial (RCT), our primary objective was to compare hydroxychloroquine (HCQ), nitazoxanide (NIT) and ivermectin (IVE) through a randomized prospective open-label observational study to detect superiority or non-inferiority, from which we would choose one drug to compare with antiandrogens, including spironolactone (SPIRO), dutasteride (DUTA) and proxalutamide (PROXA), that were our primary hypothesis as possibly being the target to actually demonstrate antiviral effects, with consequent improvements in clinical outcomes, in a placebo-control double-blind randomized clinical trial (RCT) (47-49).

However, during the observational study, we statistically noticed that besides disclosing similar clinical outcomes between them, HCQ, NIT and IVE apparently demonstrated better outcomes than those expected for COVID-19. The unexpected apparent positive results forced us to question whether the employment of full placebo-control studies in early stage COVID-19 would still be ethically acceptable. Figure 1 summarizes the path towards the ethical questions on the use of placebo in early COVID-19 patients.

**Figure 1.**
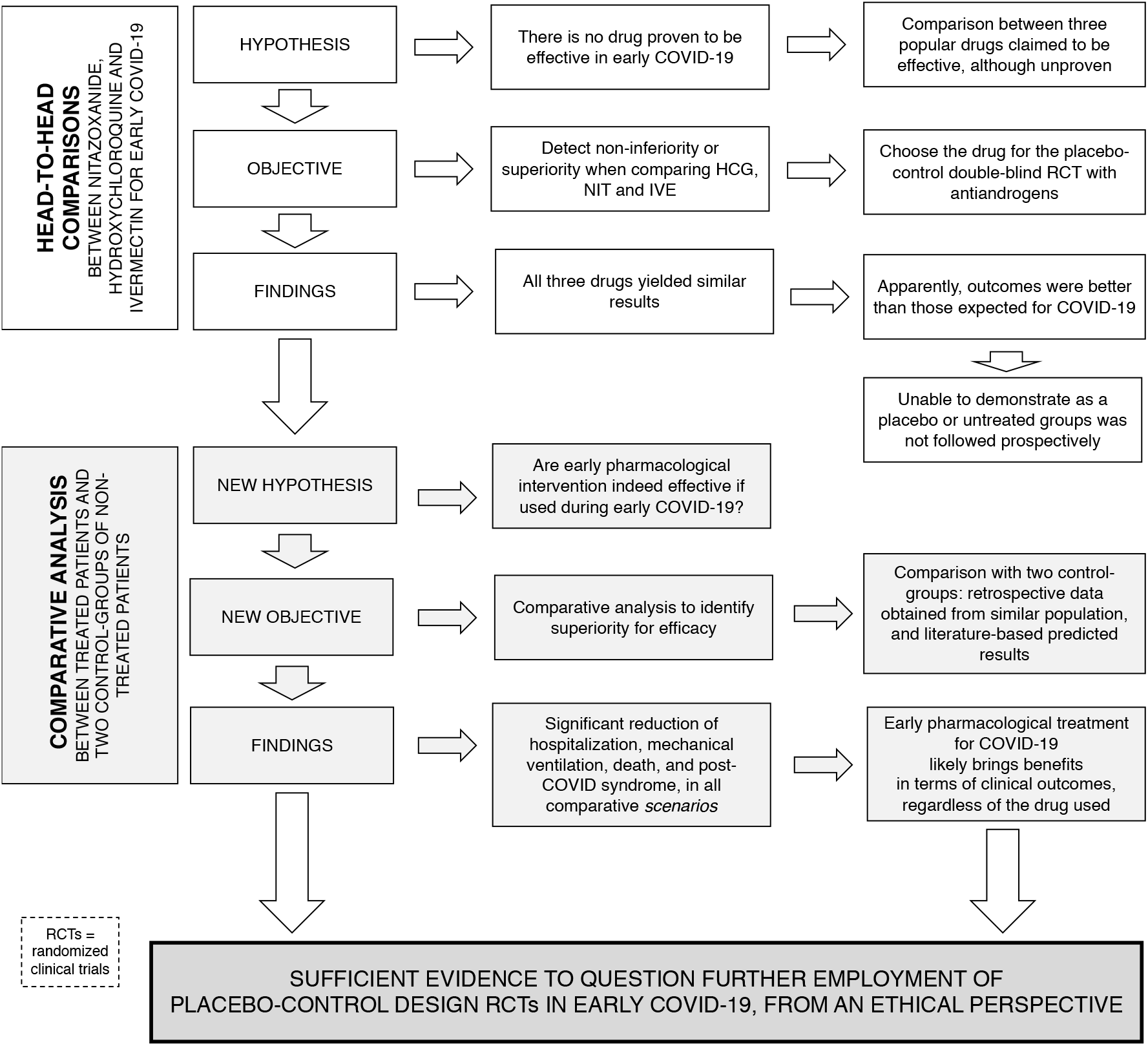
Rationale for the ethical questioning on the employment of placebo-control design in RCTs for early COVID-19.

The objective of the present study was elucidate whether the conduction of a full placebo-control RCT was still viable from an ethical perspective, through two independent comparative analysis of the present findings, with two control-groups: one of local patients with COVID-19 with similar characteristics that either refused or were not offered therapy, since the standard of care did not require early treatments during the period that these patients had COVID-19, and the other one of expected and estimated outcomes based on data generated from a thorough, structured review of the literature. We aimed to evaluate whether differences were actual, their social repercussions, and if these discrepancies were irrefutable, leading to mandatory changes in the design of further RCTs on early COVID-19, by the exclusion of placebo groups.

## Materials and methods

### Subject selection for the Active Group (AG)

Patients for the Active Group (AG) were recruited through social media, group messages and various types of medical centers located in Brasilia, Brazil, in case of suspected or confirmed COVID-19. Suspected cases for COVID-19 were defined as presenting any specific or unspecific symptom among upper respiratory tract, musculoskeletal, gastrointestinal, neurological and cardiovascular systems, from which fever, shortness of breath, anosmia or ageusia were not s*ine-quo-non* manifestations. These cases were submitted to a rtPCR-SARS-CoV-2 (Abbott RealTime SARS-CoV-2 Assay, Abbott, USA; or Cobas SARS-CoV-2, Roche, Switzerland), from which those confirmed for SARS-CoV-2 were checked for inclusion criteria and included if criteria were met. Patients previously confirmed for COVID-19 underwent directly the inclusion criteria, and were included if criteria were fulfilled.

Once patients were confirmed for COVID-19, inclusion criteria included: 1. Above 18 years old; 2. Less than seven days since the beginning of symptoms and 72 hours since the diagnosis of COVID-19; 3. Lack of use of the following drugs for more than 48 hours, including hydroxychloroquine, nitazoxanide, ivermectin or any glucocorticoid regimen, 4. Lack of clinical or radiological signs of complications related to COVID-19 or in its second or third stage. Uncompensated shortness of breath, median oxygen saturation (SatO2) lower than 92% in the last six hours and more than 50% of lungs affected in a chest computed tomography (CT) scan were criteria for exclusion and referral to a hospital.

Patients that fulfilled criteria for the observational study consented in a formal written manner exactly as determined by the Institutional Review Board (IRB) of the Ethics Committee of the National Board of Ethics Committee of the Ministry of Health, Brazil (CEP/CONEP: Parecer 4.173.074 / CAAE: 34110420.2.0000.0008). The RCT resulted from this active observational is registered at ClinicalTrials.gov (Identifier: NCT04446429, available at clinicaltrials.gov (https://clinicaltrials.gov/ct2/show/NCT04446429?term=NCT04446429&draw=2&rank=1

### Control Groups (CGs)

Two control-groups based on comparative analysis were employed, in order to find consistency and reproducibility of the data with Control Groups 1 and 2, respectively. All data was adjusted for age, sex, and presence of comorbidities.

Control Group 1 (CG1) is a group of paired untreated patients randomly obtained retrospectively from the population of the same community that had confirmed diagnosis of COVID-19 during the same period of those included in the Pre-AndroCoV Trial. i.e., that presented COVID-19 alongside patients included in the present observational study, that either refused or were not offered a treatment with any drug between nitazoxanide, hydroxychloroquine and ivermectin. Patients were enrolled from a variety of sources, including those that were provided supportive medical standard of care for COVID-19 from the same institute that conducted the observational study, and were also obtained retrospectively in a structured manner from different private medical centers that followed up patients during COVID-19, located locally, whereas those already hospitalized at the time of recruitment or referred by hospitals were excluded, in order to avoid overestimation of hospitalization and other complication rates in untreated patients.

A second control group (Control Group 2 - CG2) resulted from a precise estimation based on a thorough and structured review of articles indexed in PubMed and MEDLINE, and statements by official government agencies and specific medical societies (50-84). From data obtained, each parameter was estimated for range, median and consistency. From the most consistent values and ranges, the least negative data was employed when results were unsimilar (above 20% of discrepancy between data), or the median adjusted for one standard deviation (SD) in favor of positive outcomes was employed when results were less variable, i.e., when differences were lower than 20% for a same parameter between different studies. Particularly, when data had heterogeneity above 100% of difference between them, only the range of the data was described (*e*.*g*., if for a specific symptom, prevalence was described as being between 10% and 80%, 10%-80% was described), while the least negative value was employed for comparison purposes.

### Patient characterization

Patients were characterized for age, sex, prevalence of obesity, hypertension, type 2 diabetes mellitus, overall comorbidities rate, and adjusted accordingly. AC were additionally characterized for the presence of other 35 diseases and 20 drug classes.

### Procedures

Only patients from AC underwent early specific pharmacological approaches based on the literature or as per the Brazilian Ministry of Health, including azithromycin 500mg daily for five days for all patients, in association with one of the following: hydroxychloroquine 400mg daily for five days, nitazoxanide 500mg *twice a day* for six days, or ivermectin 0.2mg/kg/day in a single daily dose for three days. Dutasteride, spironolactone, vitamin D, vitamin C, zinc, apibaxan, rivaroxaban, enoxaparin, and glucocorticoids were added according to clinical judgement and risk for thrombosis and progression to inflammatory stage. The choice between hydroxychloroquine, nitazoxanide, and/or ivermectin, as well as the optional use dutasteride, spironolactone (if applicable) and any other drug or supplement was based on clinical judgement, availability, and individual medical history, in a *quasi-*randomized manner.

### Parameters

Major clinical course, outcomes and endpoints were measured or calculated for all groups, including viral shedding using rtPCR-SARS-CoV-2 (Abbott RealTime SARS-CoV-2 Assay, Abbott, USA; or Cobas SARS-CoV-2, Roche, Switzerland), remission of symptoms related to acute COVID-19 not including and including anosmia and ageusia, percentage of hospitalization, mechanical ventilation and deaths, and presence of mental or physical post-COVID symptoms.

Duration of positive rtPCT-SARS-CoV-2 and clinical remission not including and including anosmia and ageusia were directly obtained from patients of AG and CG1, while estimated from the extensive descriptions in the literature for CG2. Remission of CG2 was based on data from outpatients of similar age and other characteristics presenting mild disease. The median duration of dry cough, the most prolonged symptom excluding anosmia, ageusia, was the driver of the clinical duration without anosmia and ageusia, while duration of anosmia and ageusia were estimated less precisely, since there is inconsistent data regarding their duration, and they seem to vary widely among infected individuals.

Percentage of hospitalization, mechanical ventilation and deaths were directly obtained from AC and CG1, while for CG2 these parameters were determined from the proportion between rates for those aged between 40 and 49 y/o with and without comorbidities, since presence of comorbidities was associated with up to 12 times higher complication and mortality rates. We assumed a prevalence of comorbidities of 20% among patients of the present study when calculating expected outcomes in order to avoid overestimation of expected rates of hospitalization, mechanical ventilation and deaths, and consequently avoid overestimation of prevented clinical outcomes. i.e., potential benefits of early pharmacological approaches to COVID-19.

Presence of physical, mental and overall persistency of symptoms after COVID-19 remission was determined from AC and CG1, and estimated from CG2, although data on post-COVID related symptoms are still inconsistent.

Clinical course was based on percentage and median duration of each of major symptoms, median time-to-progression to more severe states, and time spent in each of these complications, including hospitalization, intensive care unit (ICU), non-invasive oxygen regimens, mechanical ventilation, pressors, and death, and was evaluated for Days 0, 1, 2, 3, 7, 14, 30 and 60. Accordingly, WHO COVID Ordinal Outcomes was employed aiming to quantify clinical status, and was evaluated in Days 0, 7, 14, 30 and 60.

Level of certainty for each parameter was described as being 0 (not certain at all) to 10 (extremely certain), according to the level of precision of the data obtained for each specific parameter, for comparative analysis purposes. CG1 and CG2 underwent pairwise comparative analysis to evaluate consistency between world data and data generated from the local untreated population.

Control Group 1 was not evaluated for symptoms due to insufficient accuracy regarding the description of each symptom for this population. Likewise, for all other parameters, only those for which data was sufficiently described, with minimally consisting findings between different studies, were included.

We avoided statistical analyses from regions with higher case fatality ratio (CFR), as those observed in Northern Italy (69), as this could artificially increase the estimation of the number of preventable COVID-19 related complications.

Patients that were initially mild or moderate and therefore eligible for the study but required further hospitalization due to progression to more severe states were referred to an emergency unit and followed together with the hospital team.

### Amplitude effect – estimated number of preventable outcomes

Considering the presence of an exceeding number of patients that likely prevented hospitalization, mechanical ventilation, death, and post-COVID syndrome in the treated group, we also calculated estimates for number of patients that were prevented to progress to any of these outcomes. In order to avoid overestimation of potential preventable outcomes we assumed that our ability to diagnose COVID-19 during earlier stages was at least twice as higher as for overall population, leading to reduction of predicted complications.

### Statistical analysis

Full raw data for AC and CG1 is available at a public repository (https://osf.io/cm4f8/). Sample size was determined based on the assumptions thatits estimate for the chi-squared test would require 80% power to detect the difference in proportions at p = 0.05, at least 95% of subjects would complete the study, and hospitalization and death rates being between 3 and 20%, and 0.3 and 2.5%, respectively. From these assumptions, we calculated a minimum of 45 and 125 patients for each subjects to detect safety and efficacy differences, respectively, that could justify its early termination.

Nonparametric ANOVA (Kruskal-Wallis) with adjusted Dunn’s test for pairwise analyses when overall *p* < 0.05, assuming that all parameters were distributed non-normally. All statistical tests were performed using XLSTAT version 22.4.1 (Microsoft, USA).

## Results

Baseline characteristics of the present study are similar between treated (AG) and untreated patients (CG1 and CG2), and similar to than those of the literature, except for the higher prevalence of obesity in CG2 versus CG1 and treated population, while prevalence was CG1 and treated population. There was numerically lower presence of hypertension and T2DM in CG2, and comorbidities in both CG1 and CG2, compared to treated population (Table 1). CG1 did not require adjustments for age, sex, obesity and other comorbidities, as they presented similar characteristics than treated population.

**Table 1.** Baseline and overall patient characterization.

| Baseline characteristics | Overall (n = 585) | Control-group 1 (CG1) – Same population controls (n=137) | Control-group 2 (CG2) – World estimated (paired for n = 585) | p-value (overall) |
| --- | --- | --- | --- | --- |
| Sex |  |  |  | n/s |
| Male | 315 (53.8%) | 77 (56.2%) | 50-65% |  |
| Female | 270 (46.2%) | 60 (43.4%) | 35-50% |  |
| Age (y/o)<br>(Mean ± SD)<br>[Median; 95%CI] | 42.4 ± 11.3<br>[42;0.9] | 44.1 ± 10.8<br>[44;1.8] | 40-55<br>[45;2.0] | n/s |
| BMI (kg/m<sup>2</sup>)<br>(Mean ± SD)<br>[Median; 95%CI] | 25.7 ± 4.6<br>[25.1;0.4] | n/a | 24-27<br>[26.0;1.5] | n/s |
| Hypertension<br>[Number (%)] | 105 (17.9%) | 22 (16.0%) | 80 (13.7%) | n/s |
| Lipid disorders<br>[Number (%)] | 173 (29.6%) | n/a | n/a | n/a |
| Type 2 diabetes mellitus (T2DM)<br>[Number (%)] | 59 (10.1%) | 11 (8.0%) | 30 (5.1%) | n/s |
| Obesity<br>[Number (%)] | 104 (17.8%) | 23 (16.8%) | 177 (30.3%) | $< 0.0001$<br>($p =$ n/s vs CG1<br>and $< 0.0001$ vs CG2) |
| Overall comorbidities (except obesity) | 151 (25.8%) | 26 (21.2%) | 117 (20%) | n/a |
| and lipid disorders)<br>[Number (%)] |  |  |  |  |
| Angiotensin converter inhibitors (ACEi)<br>[Number (%)] | 34 (5.8%) | n/a | n/a | n/a |
| Angiotensin-2 receptor blockers (ARB)<br>[Number (%)] | 86 (14.7%) | n/a | n/a | n/a |
| Statins<br>[Number (%)] | 151 (25.8%) | n/a | n/a | n/a |
| Metformin<br>[Number (%)] | 114 (19.5%) | n/a | n/a | n/a |
| Selective serotonin reuptaker inhibitors (SSRIs)<br>[Number (%)] | 74 (12.6%) | n/a | n/a | n/a |

Table 2 describes the prevalence of major symptoms in COVID-19. Fever, shortness of breath, dry cough and fatigue were significantly less present in the participants of the pre-AndroCoV Trial compared to general population (CG2), while diarrhea and headache were more present in the present study. There was insufficient accurate data on age-adjusted prevalence of “feverish”, rhinorrhea, self-reported “sinusitis”, dizziness, weakness, arthralgia, thoracic, upper back and lower back pain, vomiting alone, conjunctival hyperemia, pre-orbital pain, and dry eyes and mouth.

**Table 2.** COVID-19 clinical characterization

| Clinical clustering | Overall (n = 585) | Control-group 2 (CG2) – World estimated (paired for n = 585) | p-value (overall) |
| --- | --- | --- | --- |
| Fever<br>*Number (%)* | 67 (11.4%) | 252 (43%) | < 0.0001 |
| “Feverish”<br>*Number (%)* | 132 (22.6%) | n/a | n/a |
| Rhinorrhea<br>*Number (%)* | 182 (31.1%) | n/a | n/a |
| Headache<br>*Number (%)* | 248 (42.4%) | 199 (34%) | 0.077 |
| Shortness of breath<br>*Number (%)* | 36 (6.1%) | 170 (29%) | < 0.0001 |
| Anosmia<br>*Number (%)* | 409 (69.9%) | 410 (70%) | n/s |
| Ageusia<br>*Number (%)* | 375 (64.1%) | 380 (65%)<br>[45-70%] | n/s |
| Dry cough<br>*Number (%)* | 142 (24.3%) | 398 (68%)<br>(50-83%) | < 0.0001 |
| “Sinusitis”<br>*Number (%)* | 38 (6.5%) | n/a | n/a |
| “Sore throat”<br>*Number (%)* | 117 (20.0%) | 117 (20%)<br>(5-61%) | n/s |
| Dizziness<br>*Number (%)* | 113 (19.3%) | n/a | n/a |
| Fatigue<br>*Number (%)* | 201 (34.5%) | 439 (75%) | < 0.0001 |
| Weakness<br>*Number (%)* | 167 (28.5%) | n/a | n/a |
| Myalgia<br>*Number (%)* | 184 (31.4%) | 210 (36%) | n/s |
| Arthralgia<br>*Number (%)* | 57 (9.7%) | n/a | n/a |
| Thoracic pain<br>*Number (%)* | 169 (28.9%) | n/a | n/a |
| Upper back<br>pain<br>*Number (%)* | 72 (12.3%) | n/a | n/a |
| Lower back<br>pain<br>*Number (%)* | 23 (3.9%) | n/a | n/a |
| Diarrhea<br>*Number (%)* | 131 (22.4%) | 76 (13%) | 0.015 |
| Nauseas<br>*Number (%)* | 40 (6.8%) | 64 (11%) | n/s |
| Vomiting<br>*Number (%)* | 3 (0.5%) | n/a | n/a |
| Abdominal<br>pain<br>*Number (%)* | 34 (5.8%) | 29 (5%) | n/s |
| Conjunctival<br>hyperemia<br>*Number (%)* | 115 (19.6%) | n/a | n/a |
| Pre-orbital<br>pain<br>*Number (%)* | 45 (7.7%) | n/a | n/a |
| Dry eyes<br>*Number (%)* | 6 (1.0%) | n/a | n/a |
| Dry mouth<br>*Number (%)* | 3 (0.5%) | n/a | n/a |

Main clinical outcomes are summarized in Table 3. Duration of positive rtPCR-SARS-CoV-2 was 36.5% and 31.5% lower in AG compared to CG1 and CG2, respectively (p < 0.0001 for both), duration of clinical manifestations was reduced by 70% to 73% not including anosmia and ageusia, and 70% to 85% including both (p < 0.0001 for both), and demonstrated ability to prevent between 140 and 197 hospitalizations, 50 to 66 patients needing mechanical ventilation, five to 14 deaths, and 404 to 875 patients persisting with post-COVID symptoms, for every 1,000 cases (< 0.0001, except for mechanical ventilation and death when unadjusted, although < 0.0001 when adjusted for population models).

**Table 3.**
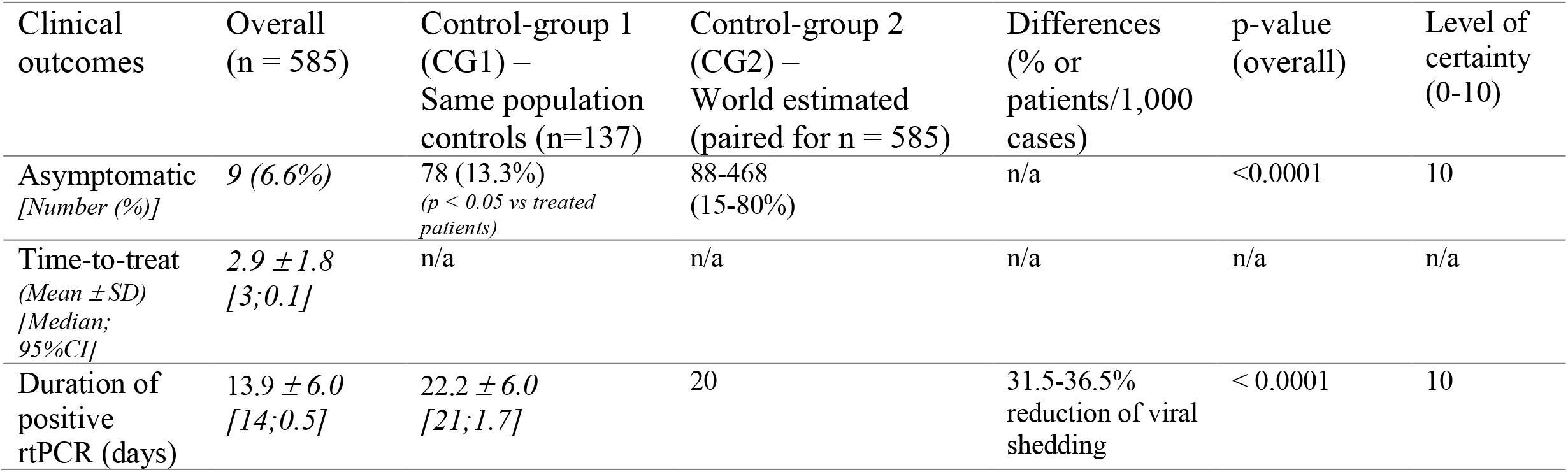
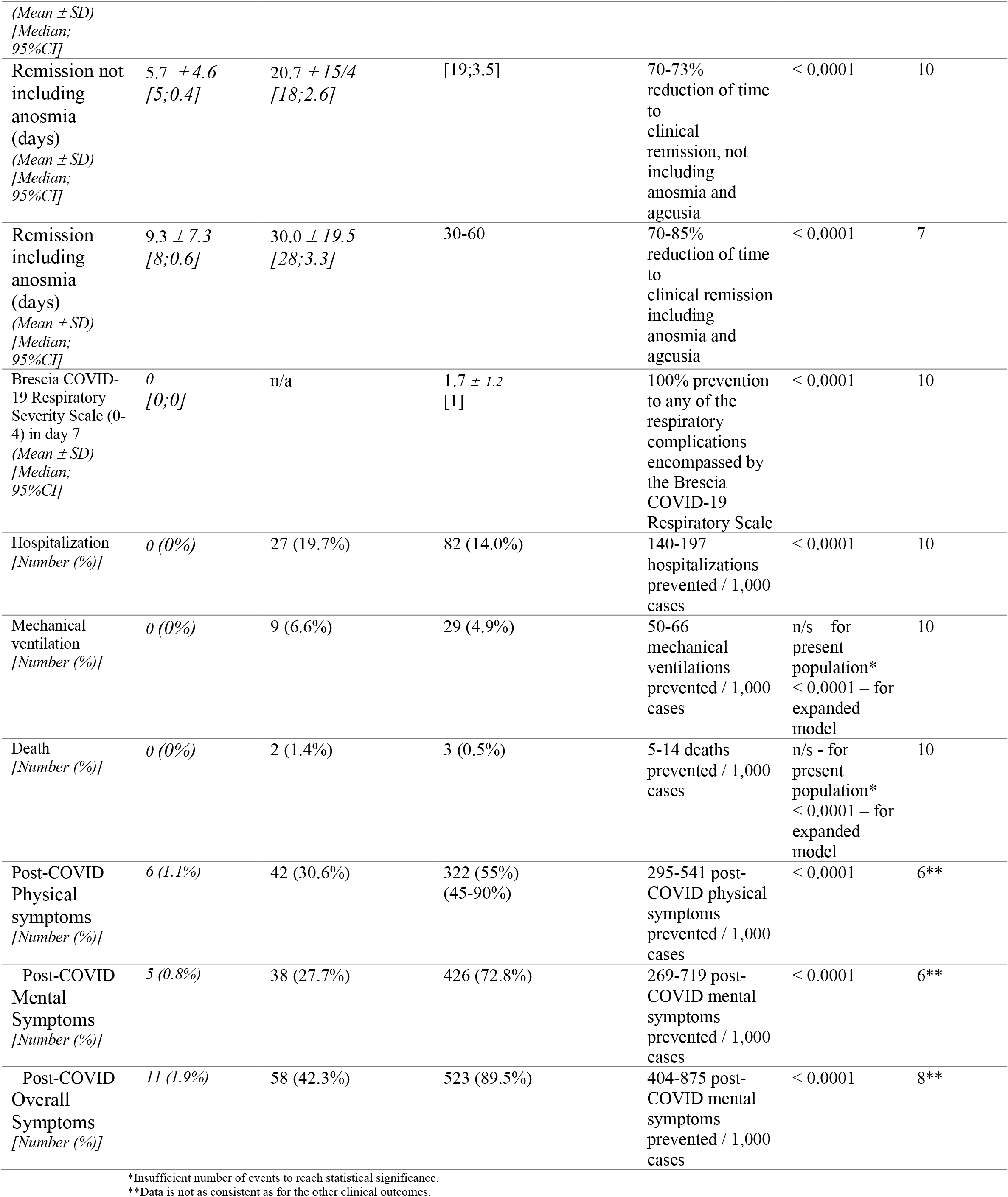
Clinical outcomes.

| Clinical<br>outcomes | Overall<br>(n = 585) | Control-group 1<br>(CG1) –<br>Same population<br>controls (n=137) | Control-group 2<br>(CG2) –<br>World estimated<br>(paired for n = 585) | Differences<br>(% or<br>patients/1,000<br>cases) | p-value<br>(overall) | Level of<br>certainty<br>(0-10) |
| --- | --- | --- | --- | --- | --- | --- |
| Asymptomatic<br>*[Number (%)]* | 9 (6.6%) | 78 (13.3%)<br>*(p < 0.05 vs treated*<br>*patients)* | 88-468<br>(15-80%) | n/a | <0.0001 | 10 |
| Time-to-treat<br>*(Mean ± SD)*<br>*[Median;*<br>*95%CI]* | 2.9 ± 1.8<br>[3;0.1] | n/a | n/a | n/a | n/a | n/a |
| Duration of<br>positive<br>rtPCR (days) | 13.9 ± 6.0<br>[14;0.5] | 22.2 ± 6.0<br>[21;1.7] | 20 | 31.5-36.5%<br>reduction of viral<br>shedding | < 0.0001 | 10 |
| *(Mean ± SD)*<br>*[Median;*<br>*95%CI]* |  |  |  |  |  |  |
| Remission not including anosmia (days)<br>*(Mean ± SD)*<br>*[Median;*<br>*95%CI]* | 5.7 ± 4.6<br>*[5;0.4]* | 20.7 ± 15/4<br>*[18;2.6]* | [19;3.5] | 70-73% reduction of time to clinical remission, not including anosmia and ageusia | < 0.0001 | 10 |
| Remission including anosmia (days)<br>*(Mean ± SD)*<br>*[Median;*<br>*95%CI]* | 9.3 ± 7.3<br>*[8;0.6]* | 30.0 ± 19.5<br>*[28;3.3]* | 30-60 | 70-85% reduction of time to clinical remission including anosmia and ageusia | < 0.0001 | 7 |
| Brescia COVID-19 Respiratory Severity Scale (0-4) in day 7<br>*(Mean ± SD)*<br>*[Median;*<br>*95%CI]* | 0<br>*[0;0]* | n/a | 1.7 ± 1.2<br>*[1]* | 100% prevention to any of the respiratory complications encompassed by the Brescia COVID-19 Respiratory Scale | < 0.0001 | 10 |
| Hospitalization<br>*[Number (%)]* | 0 (0%) | 27 (19.7%) | 82 (14.0%) | 140-197 hospitalizations prevented / 1,000 cases | < 0.0001 | 10 |
| Mechanical ventilation<br>*[Number (%)]* | 0 (0%) | 9 (6.6%) | 29 (4.9%) | 50-66 mechanical ventilations prevented / 1,000 cases | n/s – for present population*<br>< 0.0001 – for expanded model | 10 |
| Death<br>*[Number (%)]* | 0 (0%) | 2 (1.4%) | 3 (0.5%) | 5-14 deaths prevented / 1,000 cases | n/s - for present population*<br>< 0.0001 – for expanded model | 10 |
| Post-COVID Physical symptoms<br>*[Number (%)]* | 6 (1.1%) | 42 (30.6%) | 322 (55%)<br>(45-90%) | 295-541 post-COVID physical symptoms prevented / 1,000 cases | < 0.0001 | 6** |
| Post-COVID Mental Symptoms<br>*[Number (%)]* | 5 (0.8%) | 38 (27.7%) | 426 (72.8%) | 269-719 post-COVID mental symptoms prevented / 1,000 cases | < 0.0001 | 6** |
| Post-COVID Overall Symptoms<br>*[Number (%)]* | 11 (1.9%) | 58 (42.3%) | 523 (89.5%) | 404-875 post-COVID mental symptoms prevented / 1,000 cases | < 0.0001 | 8** |

\*Insufficient number of events to reach statistical significance.
\*\*Data is not as consistent as for the other clinical outcomes.

Regarding clinical course of COVID-19, Days 1, 2, 3, 7 and 14 showed significantly more rapid improvement rates in the population treated in the pre-AndroCoV Trial than CG1 and CG2 (p < 0.0001, except for Day 14 – p – 0.033) (Table 4). In the WHO COVID Ordinal Outcomes (Table 5), treated patients were significantly better than untreated (CG1 and CG2) in Days 7, 14 and 30.

**Table 4.** Clinical course.

| Clinical course | Overall females (n = 270) | Overall males (n = 315) | Overall (n = 585) | Control-group 1 (CG1) – Same population controls (n=137) | Control-group 2 (CG2) – World estimated (paired for n = 585) | p-value (overall) | Level of certainty |
| --- | --- | --- | --- | --- | --- | --- | --- |
| **Days -7 to -4**<br>*Median; 95%CI*<br>(% asymptomatic)<br>(pairwise p-value) | 100 (4.3)<br>(75.9% asymptomatic) | 100 (3.3)<br>(84.1% asymptomatic) | 100 (2.7)<br>[470 (80.3%)]<br>asymptomatic | n/a | n/a | n/a | n/a |
| **Days -3 to -1**<br>*Median; 95%CI*<br>(% asymptomatic)<br>(pairwise p-value) | 10 (4.8)<br>(24.1% asymptomatic) | 10 (4.7)<br>(30.5% asymptomatic) | 10 (3.4)<br>[161 (27.5%)]<br>asymptomatic | n/a | n/a | n/a | n/a |
| **Day 0**<br>*Median; 95%CI*<br>(% asymptomatic)<br>(pairwise p-value) | 20 (4.2)<br>(12.6% asymptomatic) | 10 (4.3)<br>(18.3% asymptomatic) | 20 (3.0)<br>[93 (15.9%)]<br>asymptomatic | [10] | [20] | n/s | 10 |
| **Day 1**<br>*Median; 95%CI*<br>(% asymptomatic)<br>(pairwise p-value) | 60 (4.0)<br>(12.6% asymptomatic) | 50 (4.0)<br>(19.4% asymptomatic) | 50 (2.9)<br>[95 (16.2%)]<br>asymptomatic | [0] | [10] | < 0.0001 | 3 |
| **Day 2**<br>*Median; 95%CI*<br>(% asymptomatic)<br>(pairwise p-value) | 90 (3.2)<br>(20.4% asymptomatic) | 90 (3.4)<br>(25.4% asymptomatic) | 90 (2.4)<br>[135 (23.1%)]<br>asymptomatic | [10] | [20] | < 0.0001 | 3 |
| **Day 3**<br>*(Mean ± SD)*<br>*[Median; 95%CI]*<br>(pairwise p-value) | 95 (2.7)<br>(36.3% asymptomatic) | 90 (2.6)<br>(35.9% asymptomatic) | 90 (1.9)<br>[211 (36.1%)]<br>asymptomatic | [20] | [30] | < 0.0001 | 4 |
| **Day 7**<br>*Median; 95%CI*<br>(% asymptomatic)<br>(pairwise p-value) | 100 (1.7)<br>(66.3% asymptomatic) | 100 (1.2)<br>(72.7% asymptomatic) | 100 (1.0)<br>[408 (69.7%)]<br>asymptomatic | [50] | [60] | < 0.0001 | 7 |
| **Day 14**<br>*Median; 95%CI*<br>(% asymptomatic)<br>(pairwise p-value) | 100 (0.9)<br>(90.0% asymptomatic) | 100 (0.5)<br>(92.4% asymptomatic) | 100 (0.5)<br>[534 (91.3%)]<br>asymptomatic | [80] | [90] | 0.033 | 8 |
| **Day 21**<br>*Median; 95%CI*<br>(% asymptomatic) | 100 (0.7)<br>(97.4% asymptomatic) | 100 (0.1)<br>(92.8% asymptomatic) | 100 (0.4)<br>[568 (97.1%)]<br>asymptomatic | [90] | [100] | n/s | 4 |
| (% asymptomatic)<br>(pairwise p-value) |  |  |  |  |  |  |  |
| Day 30<br>(Median; 95%CI)<br>(% asymptomatic)<br>(pairwise p-value) | 100 (0.4)<br>(98.5%)<br>asymptomatic) | 100 (0.1)<br>[190 (99.0%)<br>asymptomatic] | 100 (0.2)<br>[577 (98.6%)<br>asymptomatic] | [100] | [100] | n/s | 8 |
| Day 60<br>(Mean ± SD)<br>[Median; 95%CI]<br>(pairwise p-value) | 100 (0.0)<br>(100%)<br>asymptomatic) | 100 (0.0)<br>(100%)<br>asymptomatic) | 100 (0.0)<br>[585 (100%)<br>asymptomatic] | [100] | [100] | n/s | 8 |

**Table 5.**
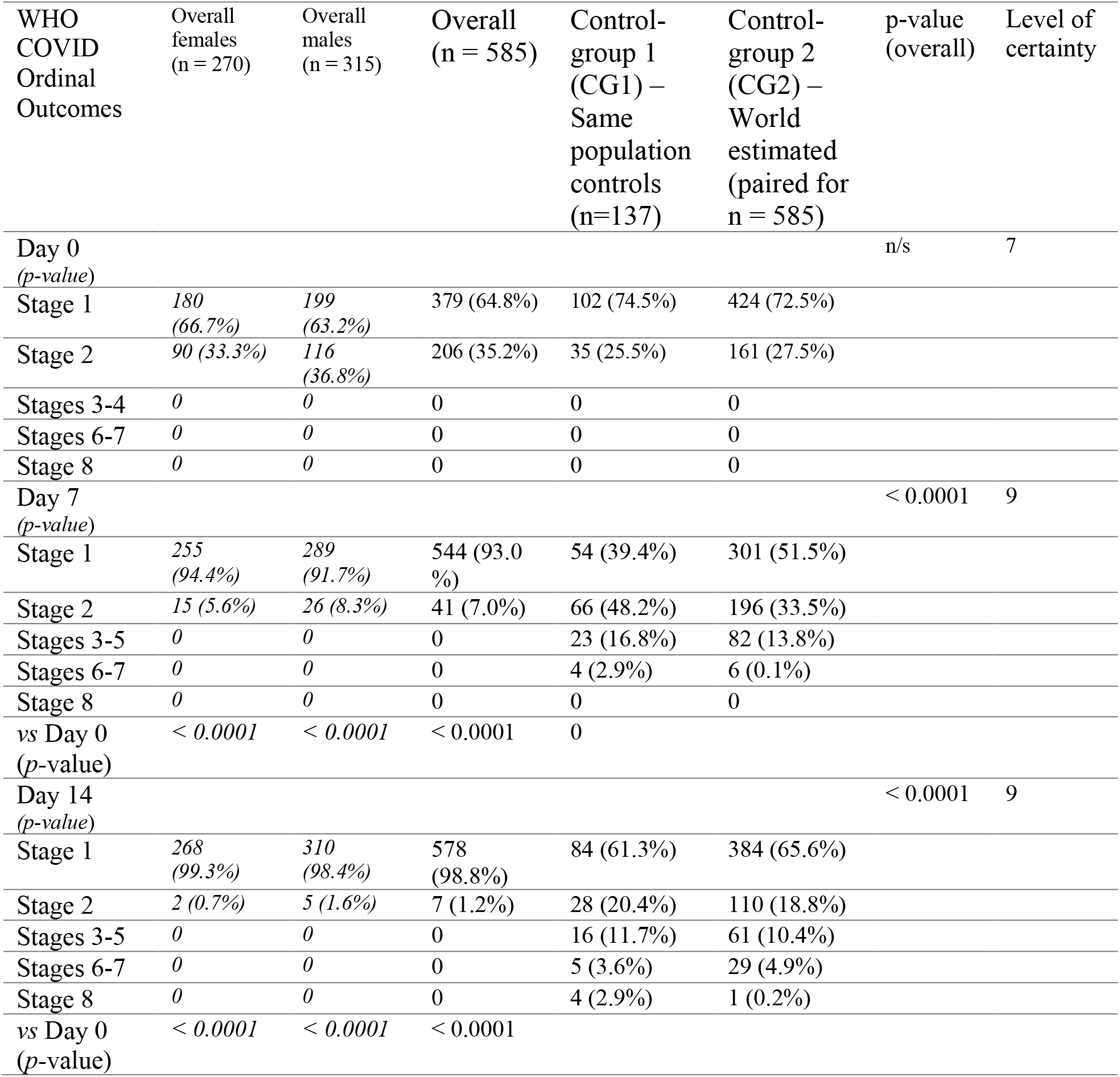
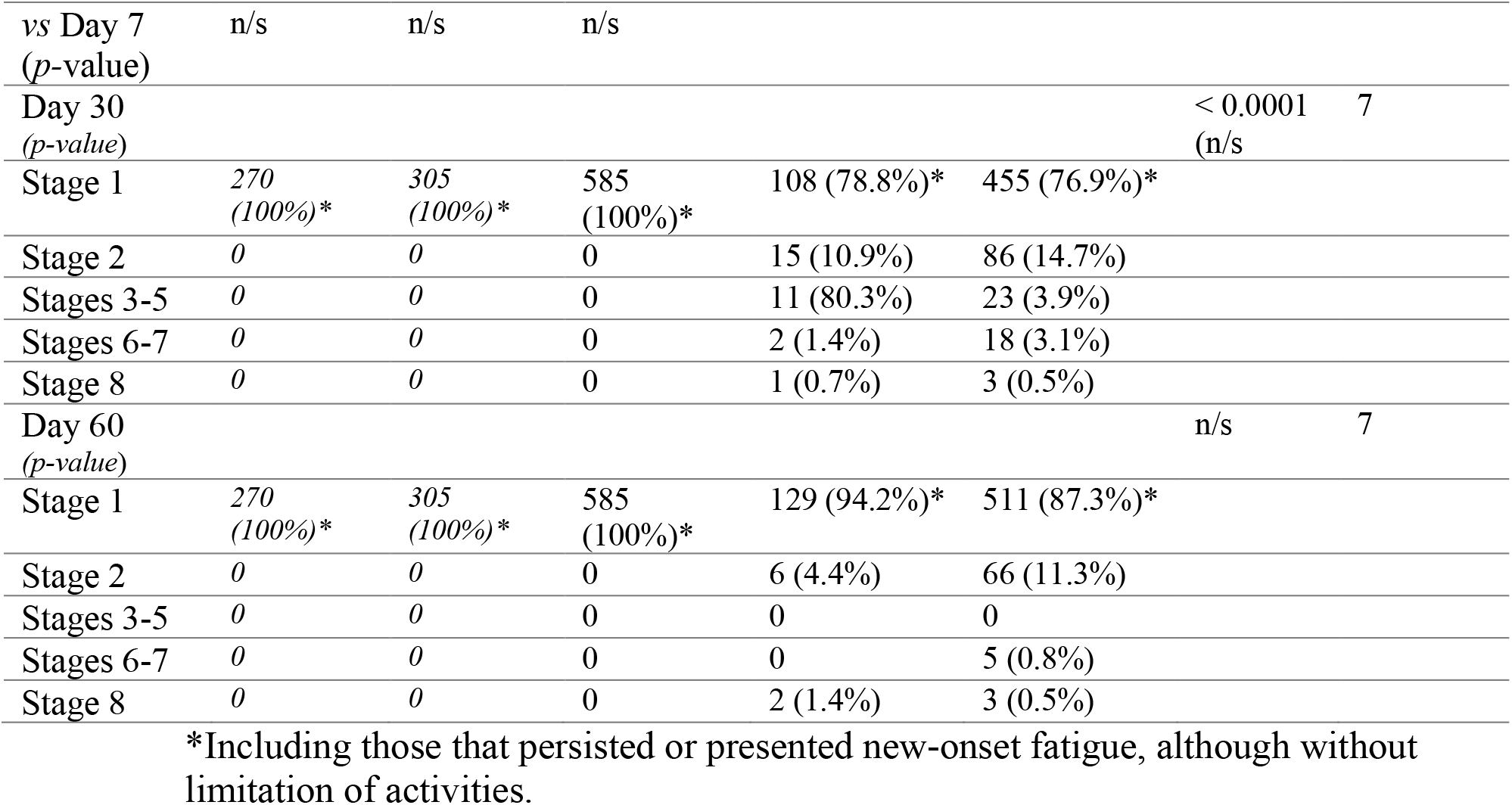
WHO COVID Ordinal Outcomes.

CG1 and CG2 presented similar characteristics and disease parameters, except for obesity prevalence (CG1 > CG2) and slightly lower number of patients in Stage 1 on Day 7 of the WHO COVID Ordinary Outcomes.

## Discussion

### Superiority of early pharmacological interventions for COVID-19: apparent or actual?

When we conducted the observational study comparing HCQ, NIT and IVE, we presumed that there were no actual effective options for early COVID-19, i.e., that none of the drugs would confer any protection. As per the design, we did not include patients that did not undergo any specific treatment, since our primary objective was to perform a *head-to-head* comparison. However, once none of the patients were hospitalized, needed mechanical ventilation, or deceased, questions regarding the superiority of any treatment over none were raised, but could be answered within the study per se, as we did not originally include untreated patients, precluding us from any conclusion regarding the overall efficacy.

When we detected an apparent superiority in terms of efficacy for HCQ, NIT and IVE, this was still speculative, and was not the primary endpoint of the open-label randomized prospective observational study. To respond to this question raised by the clear differences between our treated and overall untreated populations, we performed the present comparative analysis based in two different control-groups, in order to detect reproducibility and consistency between both comparisons.

However, the shortage of a thorough characterization of clinical manifestations during early COVID-19 due to a large variability of symptoms and inability to detect COVID-19 in the first days of symptoms, and the substantial variability in the prevalence of symptoms reported, possibly due to methodological differences in the search for manifestations was a barrier for a precise comparative analysis.

Despite the limitations for a precise characterization of early COVID-19, associated with the fact that for ethical reasons differences were underestimated, the present comparative analysis revealed differences unlikely to be random for the most relevant clinical outcomes, with potential ability to prevent a non-negligible number of hospitalizations, mechanical ventilations, and deaths. The analysis disclosed approximate reductions of one third in viral shedding, two thirds in clinical duration, and 100% prevention of hospitalization, mechanical ventilation, and deaths, when compared to both CG-1 and CG-2, together and separately, which were shown to represent substantial and possibly conclusive improvements regarding the benefits, when using one of the three pharmacological interventions in early stage COVID-19, at least in when combined with azithromycin, and in majority of the cases, zinc, vitamin C and vitamin D.

Since the median age was below 60 y/o and the prevalence of less than 20% and 30% of obesity and comorbidities, respectively, and COVID-19 related complications are highly correlated with risk factors, in an exponential manner with aging, the ability to detect differences in lower risk populations allow us hypothesize that in higher-risk groups of patients infected by COVID-19 would potentially lead to more substantial differences due to the larger number of complication events, allowing more visible distinctions between groups, although this is yet to be determined.

Besides the consistent findings between comparisons of AG with CG1 and CG2, CG1 and CG2 presented similar results for virtually all parameters and outcomes, providing further consistency and strength in the findings and estimates for untreated patients.

### Avoiding overestimation of benefits from early pharmacological approaches to COVID-19

One challenge when comparing research data with external or retrospective analysis of populations, not initially included and not encompassed by the primary objectives, is to avoid authors’ bias of overestimating their findings, in particular in the context of the demonstration of efficacy of potential major public and social relevance. To overcome this potential bias, we purposely adjusted all parameters towards to underestimation of risks, complications and negative data on untreated populations, and underestimation of benefits among those treated for COVID-19.

Two major tools were employed to avoid overestimation of our findings: 1. The use of two independent control-groups, one of similar population, and the other one based on the literature, and 2. Extrapolative underestimation of COVID-19 negative outcomes and data.

The CG1 is based on a local population with similar age, proportion between sex, ethnic, socioeconomic and cultural characteristics, aiming to avoid differences due to health disparities.

For the estimation of the CG2, we have considered slightly lower disease duration, hospitalization, mechanical ventilation, death and post-COVID rates than those described by the literature. In particular, we avoided the use of studies that included hospitalized patients, even for evaluation of disease duration, even in a proportion of patients that were expected to be hospitalized, since we tended to underestimate the risks of untreated and overestimate our negative findings.

Although patients from the AG were detected in the early COVID-19 stage, they cannot be considered as non-critical, but uncertain cases. However, although patients had uncertain clinical course, for comparative purposes we avoided the use of data from critical untreated patients, and from non-critical cases instead, leading to lower negative outcomes in the estimation of untreated patients. In addition, while median age of the present population corresponded to the median age of majority of studies with hospitalized patients, we excluded studies that had higher median age since these studies could overestimate risks related to COVID-19 inherent to aging.

Noteworthy, mortality rates below 0.5% were only obtained through the analysis of seroprevalence compared to number of deaths in a specific population at a specific time, which concurrently demonstrates that asymptomatic patients represent over 80% of those infected by COVID-19. Since our study only had fewer than 15% of patients being asymptomatic, our comparisons with mortality rates should be based on CFR of populations that presented positive rtPCR-SARS-CoV-2 due to symptoms or direct contact with confirmed COVID-19 cases.

### Particularities of the present study

Some specificities of the pre-AndroCoV Trial may have contributed for the characteristic profile of the studied population and notably improved outcomes.

Possibly because we have actively searched for comorbidities that could influence risks in COVID-19, our prevalence of comorbidities was higher compared to sex- and age-matched untreated populations, even with lower BMI (when compared to CG2, but no CG1), which could have negatively influenced outcomes in the AG, although underdiagnosis of comorbidities in CG2 is possible.

The potential protectives role of statins, metformin, ACEi and ARB for COVID-19, which was regularly used by approximately 30% of the AG, may have contributed to milder COVID-19 presentation in the AG.

In addition, the non-mandatory presence of fever, shortness of breath or cough to be considered as suspected for COVID-19 may have apparently changed the profile of clinical manifestations. The active search for symptoms resulted in higher prevalence of anosmia and ageusia when compared to literature

Whether and until which extent the change in COVID-19 detection towards a more sensitive diagnosis may have affected outcomes in a positive manner is unknown, but possible. Correspondingly, a more aggressive approach to the patient suspected for COVID-19 may have been crucial for the better outcomes found in the AG, when compared CGs.

### Post-COVID syndrome as an outcome

While mortality plays a key outcome in COVID-19, the notorious presence of persisting symptoms after COVID-19 remission has called attention to the chronic aspects, possibly mediated by the triggering of immunologic *maladaptations*. Persistent fatigue, brain fog, reduction of cognitive functions, impaired muscle recovery, decreased physical capacity, reduced fertility and sexual function, and psychiatric manifestations not fully justified by post-traumatic stress disorder (PTSD), with substantial similarities with Chronic Fatigue Syndrome (CFS) and Burnout syndrome, are among the most commonly described symptoms, and may affect up to 85% of patients (85-93). Because of the potential long-term impairment of life quality, prevention of post-COVID symptoms should be considered as a major endpoint when approaching COVID-19.

In the present study, differences in the prevalence of post-COVID symptoms between treated and untreated populations were larger than differences in any other parameter. This finding must be emphasized as an additional benefit that may overcome potential risks of the drugs uses *per se, i*.*e*., even in a hypothetical absence of other benefits, prevention of post-COVID syndrome could be alone sufficient to justify early pharmacological approaches to COVID-19.

### Estimates of the impact of early pharmacological approaches in COVID-19 clinical outcomes

Estimates for the impact of early pharmacological treatment for COVID-19 have been overall underestimated in the present analysis, in order to prevent identification of not actual benefits.

In all *scenarios*, reduction of deaths and long-term consequences were meaningful, specially when analyzed through a public health perspective. At least 140,000 hospitalizations, 5,000 deaths and 250,000 long-term persistence of symptomatic subjects could be prevented for every 1,000,000 cases treated before seven days of symptoms.

The social and human relevance of these findings are of major importance in terms of absolute number of prevented complications, while risks related to the drugs proposed are largely overcome by benefits, particularly in a pandemic with massive number of cases.

### The decision of no longer use of placebo in early COVID-19

Although the employment of placebo-control study design was initially accepted due to the uncertain risks of development of complications from COVID-19. ethical issues in the employment of placebo-control studies on COVID-19 were raised after the overwhelming differences in terms of results. The prevention of hospitalization and mechanical ventilation in approximately 14% and 5% of patients affected by symptomatic COVID-19, and the reduction of 90 to 95% of patients developing post-COVID impairment of life quality are arguments initially against the continuation of full placebo-control studies for COVID-19, at least in its early stages.

There is not such a specific point from which it becomes ethically questionable to continue a clinical trial. However, the fact that a placebo control is necessary to demonstrate efficacy is not sufficient to justify it employment in all circumstances. While without the knowledge from placebo-control studies it becomes harder to obtain efficacy data, for life-threatening conditions, from the perspectives of both percentage and absolute number of affected patients, it would not be ethical to ask participants to accept known risks in case an unproven but possible and largely safe option could be offered, in a manner that risks of postponing treatment is not negligible. In summary, the use of safe options that although unproven demonstrate plausibility and preliminary observational positive results is highly recommended in the absence of established effective treatments.

When analyzing the use of placebo, the peculiarities of the COVID-19 pandemic should be highlighted, in particular regarding the *social value* of early treatment, once superiority has become evident. In the present particular case, one point that must be considered is the fact that drugs used for COVID-19 in the present study have solid safety profile and virtual no risk of serious adverse effects or complications. In common, hydroxychloroquine, nitazoxanide and ivermectin have been used for a wide range of infectious and non-infectious diseases in the long-term, with apparent steady safe profile and lack of overwhelming risks, when used in large populations, with favorable cost-effectiveness, even when used as preventive approaches to low-risk diseases, which reinforce their safety. Considering that all three molecules have sufficient safety to be used for lower-risk disorders, even in a preventive basis, it seems intuitive that their use for early COVID-19, when antiviral approaches tend to be more efficient, would be recommended, at least until evidence shows otherwise, since risks of complications, death and chronic symptoms caused by COVID-19 are known, plausibility and preliminary data exist, and safety is well-established for all three molecules. Also, in the present study, outcomes between the three drugs were similar, revealing a non-inferiority of any of them in terms of efficacy. The lack of severe adverse effects and complications also reinforced their safety profile for COVID-19.

Since respiratory state can decompensate very rapidly, besides being following closely, the timing to intervention is critical, and early pharmacological approaches showed to be likely efficient to prevent acute respiratory insufficiency.

The concept of *clinical equipoise*, although controversial and not universally accepted, finds its best example on early pharmacological approaches for COVID-19 (100-105). First, it is genuinely uncertain whether this modality could lead to improved outcomes. Second, the spread off-label use performed before initially required double-blind placebo-control RCTs. To overcome ethical dilemmas, the initial purpose of performing a comparative analysis between hydroxychloroquine, nitazoxanide and ivermectin was to determine whether one of these options would demonstrate superior efficacy over others, and then employ the drug as a full placebo-control double-blind RCT together with an antiandrogen (spironolactone, dutasteride and proxalutamide). Hence, null hypothesis was inherently considered in the beginning.

The lack of a specific threshold of evidence of superiority had not been established in the beginning, since the primary objective was to determine which of the three drugs could possibly demonstrate better efficacy while maintaining safety, in addition to the fact that COVID-19 still had uncertain estimates of complication rates by the time of the design of the present study, precluding an accurate description of stopping guidelines. With the unexpected results of lack of progression of lung injury, hospitalization, mechanical ventilation, death, and as longitudinally analyzed, post-COVID symptoms, the actual uncertainty as part of the clinical equipoise has clearly become no longer uncertain, but highly likely instead. Although Interim analysis already precluded investigators from performing a full placebo-control in the RCT with sufficient evidence with 390 patients, the bias of investigators as overestimating the benefits of proposed intervention and the fact that for some clinical *equipoise* requires overall medical community to be convinced that findings are consistent, reproduceable and unquestionable, we maintained until we reached 585 patients, with consistent results throughout the study.

In the case of the pre-AndroCoV-Trial, research ethics applied to the COVID-19 pandemic, the extreme *social value* attributed to the unexpected superiority detected in any pharmacological approach, versus those not treated, the amplitude effects of the findings, and the estimates on the impacts of the changes towards earlier detection and prompt pharmacological intervention for COVID-19 allowed us determine that placebo was no longer needed, and could actually start to affect the therapeutic obligation of medical providers to offer the now demonstrated efficient therapies for those diagnosed with COVID-19 until seven days of symptoms. The superiority of the treatment for COVID-19 has become clearly established in the accruing of comparative treatment results. Although boundaries for superiority in terms of efficacy tend to be stricter than those for safety, the superiority has become evident.

As mentioned before, since the present findings were obtained with lower risk populations, which requires more prominent differences and larger samples, in order to the superiority in terms of efficacy, the potential benefits for populations of higher risk will likely be amplified.

Since COVID-19 is of public health importance, and impact of the present findings is likely large, we were no longer ethically allowed to start or continue placebo-control RCTs, and we found mandatory to communicate our findings to overall scientific community.

In summary, the three proposed drugs have strong safety profile, have been long used for a variety of diseases, have low treatment costs, and may benefit a massive number of subjects, including the reduction from 60 to 80% of prevalence of prolonged symptoms after COVID-19 remission to 2.5 to 5% observed in treated patients, use of placebo for further studies would become harder to be ethically justified. In addition, the presence of three options allow prescribes to choose, according to medical judgement and perceptions over safety and efficacy. For the AndroCoV Trial, we opted for the use of nitazoxanide due to its stronger in vitro evidence, evidence to be effective against other viruses in humans, and less disputed safety questions. Figure 2 summarizes the rationale for the conclusions from the present analysis.

**Figure 2.**
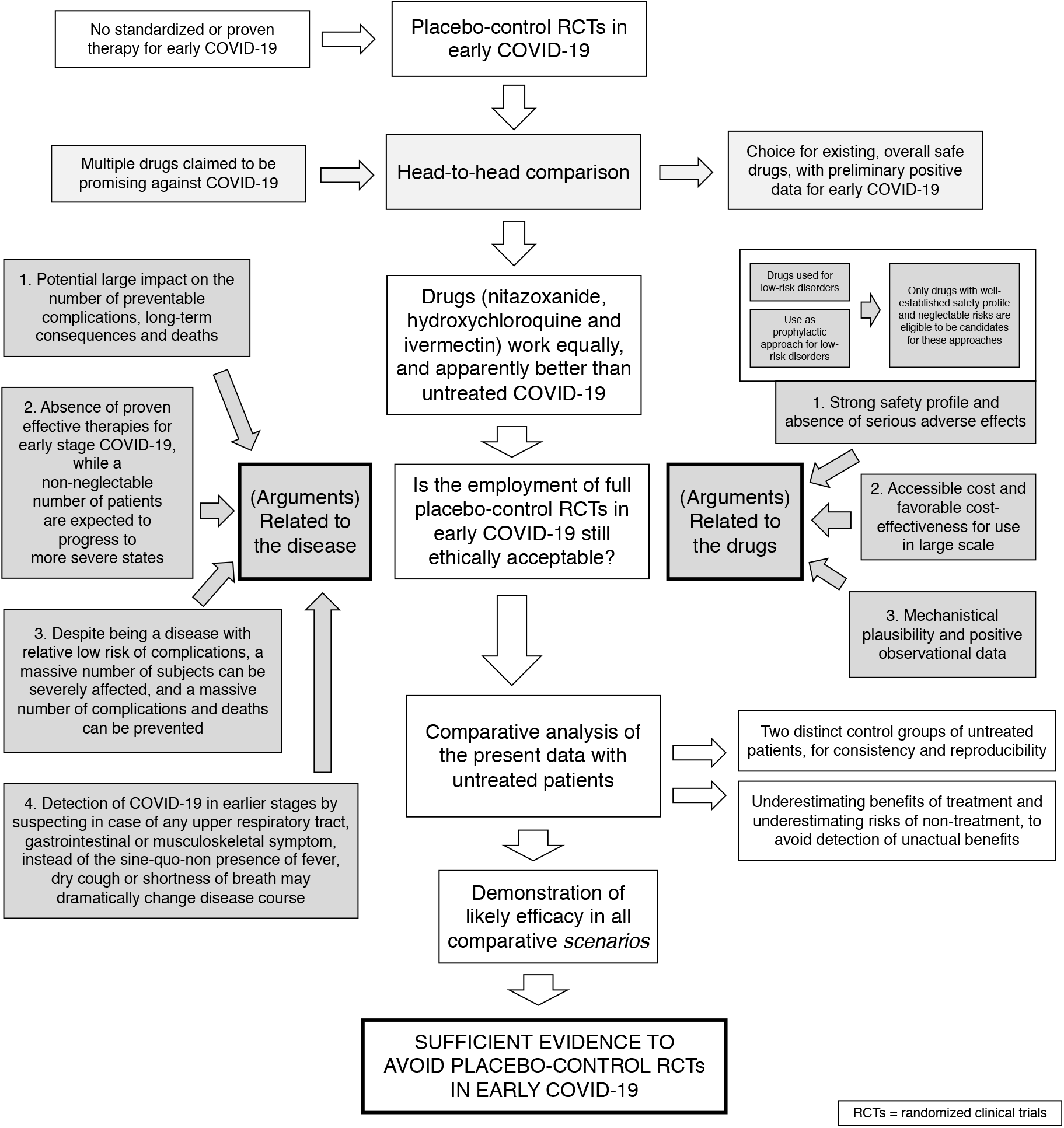
Ethics in the employment of full placebo-control RCTs in early COVID-19.

### Limitations

This is a *post-hoc* comparative analysis, combined with a comparison with two control-groups, one obtained retrospectively and one estimated and expected outcomes for the population treated for COVID-19, which may only offer evidence in case of the existence of overwhelming data to justify, while several biases should be considered when analyzing the present study.

However, the most limiting aspect of the present analysis is to reproduce the quality of patient recruitment, employment of a sensitive COVID-19 case-detection basis, turnaround time for rtPCR-SARS-CoV-2, prompt availability to provide pharmacological intervention, and, above all, availability of health professionals, essay kits, and treatments, altogether, in a worldwide basis. The feasibility to reproduce the exact same conditions of the present study in public health is uncertain, unless critical changes in policies on COVID-19 are considered. Lastly, whether and until which extent each of the aspects employed in the present analysis would impact COVID-19 outcomes alone is uncertain.

## Conclusion

Patients treated with azithromycin combined with nitazoxanide, hydroxychloroquine or ivermectin had significant reductions in viral shedding, disease duration, hospitalization, mechanical ventilation, death and post-COVID symptoms, when compared to sex-, age- and comorbidity-matched untreated patients. These differences remained when benefits of treatment and risks of COVID-19 were both underestimated. The well-established safety profile of the proposed drugs, the likely benefits and the current absence of proven therapies for early COVID-19 bring ethical questions regarding the employment of placebo-control randomized clinical trials in early COVID-19. Of the three drugs, we opted for nitazoxanide, due to more extensive demonstration of *in vitro* and *in vivo* antiviral activity, proven efficacy against other viruses in humans and steadier safety profile.

## Data Availability

Full raw data is available at a public repository (https://osf.io/cm4f8/).

